# Establishing the construct validity and Internal consistency for Urdu version of Jefferson Scale of Empathy-S (JSE-S)

**DOI:** 10.1101/2020.06.07.20114173

**Authors:** Syed Razi Muhammad

## Abstract

**BACKGROUND AND OBJECTIVES:** Empathy is an important factor in patient–physician relationship that results in better patient compliance, satisfaction, and clinical outcomes. Jefferson Scale of Empathy (JSE) assesses empathy in health professionals (HP-version) and related students (S-version). It has been translated in over 50 languages and validation studies have been performed on these translated S-version of the scale. We decided to examine empathy of Pakistani student and to study factor structure and psychometric properties of the Urdu translation of the JSE-S.

**METHODS:** 405 out of 521 students of Muhammad Medical College participated in the study. It was a cross-sectional study and the students filled in Urdu version of JSE-S questionnaire consisting of 20 items. Each item was scored on 7-points Likert scale. Total scores ranged from 20 to 140. Higher values indicated more positive attitudes toward empathic patient care, and therefore a higher degree of empathy

**RESULTS:** The mean JSE-S empathy score of the medical students was 107.22 (+/- 12.844). Cronbach’s alpha coefficient was 0.684 for the overall measure. Kaiser-Meyer-Olkin (KMO) test yielded an index of 0.764, suggesting a support for factor analysis. Bartlett’s test of sphericity was 985.518 and was highly significant (*P* = 000). A principal component analysis showed a three-factor solution that also provided support for the construct validity of the Urdu version of JSE S-version.

**CONCLUSION:** The Urdu version of JSE is a valid and reliable measure to tap empathy in a Pakistani medical student.

## Background

Empathy is defined as a cognitive attribute that engages understanding a patient’s suffering and concerns combined with an ability to communicate this understanding and an intention to help. For optimal patient outcome, strong physician-patient relationship is necessary. If patient outcome is to be rationalized, then empathy is considered as key element. Jefferson Scale of Empathy was developed by Jefferson Medical College (now Sidney Kimmel Medical College) of Thomas Jefferson University Center for Research in Medical Education and Health Care. Jefferson Scale of Empathy^2^ (JSE) is the most widely used scale to measure empathy. It is available in three versions namely

- Medical Students (S-version)
- Health Professionals (HP-version)
- Health Professional Students (HPS-version)

JSE has been translated into 56 languages. Original version is available in English which has been validated^3-5^, with permission to translate it into local language and to use it for the current research. However, Asano-Gonnella Center for research in medical education & health care did not endorse any translation and therefore it is prime responsibility of researchers to validate translated version of JSE ^6^ The translated versions of Jefferson Scale of Empathy have been used in more than 70 countries. Almost all published studies reported Cronbach’s alpha coefficients in range of 0.70 to 0.80 which indicate good reliability^7^. Yet it is important to establish construct validity and internal consistency of any translated version before it is use with confidence.

## Objective

The objective of this study is to establish the construct validity and internal consistency for Urdu translation of Jefferson Scale of Empathy Student version.

## Method

### Ethical Approval

Prior approval taken from Research and Ethical Review board of Muhammad Medical College vide letter no ERB/113/2019.

### Instrument

Urdu version of Jefferson scale of Empathy student version (JSE-S) is used. The Jefferson Scale of Empathy Student Version (JSE-S) has 20 items. Each is scored on 7-points Likert scale. Among these, 10 items are “positively worded” and scored as “Strongly Disagree=1, Strongly Agree=7”. To reduce the confounding effect of a response pattern known as “acquiescence response style, other 10 items are “negatively worded” and scored reversely as Strongly Disagree=7, Strongly Agree=1. Negatively worded items are often followed by “R”, which stands for reverse scored item. Total scores can range from 20 to 140. Higher values indicate more positive attitudes toward empathic patient care, and therefore a higher degree of empathy^5^.

As the aim of this study has been to assess the construct validity and reliability of the Urdu version of JSE-S, the average scoring of each item (with standard deviation) has been calculated and compared with those of the other studies.

Each year 100 students get admission in Muhammad Medical College, making a total of 500 students across the 5 years. As some students failed in their final examination; JSE-S Urdu version was distributed to 521 students between January 2015 and February 2015. Appropriate information and instructions given to each student to fill the questionnaire and requested to ask for clarification in case they experience difficulty in understanding any item. It was also clearly mentioned that responding survey form was not a test of their academic performance and subsequently would not be used in their grading. Participants were given choice to submit completed form anonymously. Those who mentioned names/seat number, were reassured that their identity would be kept confidential.

### Data Analysis

Survey forms with answers of 15 questions or less were considered incomplete and therefore not considered for data analysis. However when the student answered 16-19 question, the mean score of responses was calculated and this mean score was used for missing items. To calculate the adequacy and appropriateness of the data for calculating reliability analysis and construct validity, Kaiser-Meyer-Olkin (KMO) measure of sampling adequacy^8^ and the Bartlett’s Test of Sphericity^9^ were used. When data found suitable; the reliability analysis was measured using Cronbach’s α and construct validity by using principal component analysis (PCA) also known as Factor Analysis.

## Results

The breakup of 521 students with respect to study year and gender is shown in table no 1. Among 521 students, 405 students returned the form making a response rate of 77.88%.. Although only three hundreds and eighty-eight students answered all 20 items, 17 students answered between 16-19 questions, making them eligible for participation in the study. The table number 1 shows the response rate with respect to the year of study and gender. The overall mean empathy in the study is 107.22 (±12.844)/out of 140. The empathy score with ±SD is shown in table no 3. The total empathy and mean empathy of Hojat’s 3 factors are calculated, they are highest for prospective taking (6.09 ± 0.694), then compassionate care (4.97 ± 0.97), and lowest for Standing in Patient’s shoes (3.63 ±1.51) (see table no 4).

### Reliability

The internal consistency reliability of the questionnaire had a Cronbach’s alpha coefficient of 0.684 and a Cronbach’s alpha based on standardized items of 0.704.

### Cronbach’s Alpha if item deleted

An analysis was run to see how much alpha is affected if each item was deleted. Results are summarized in table 5. Deletion of items causes minimal change to alpha ranging from .650 (Item 14) to 0.697 (Item 18). From this analysis, it appears that all 20 items designed to measure JSE-S, work well and contribute to the overall reliability of JSE-S Urdu Version.

### Construct validity

To analyze that the Urdu version of JSE-S actually measures the empathy in the same way as the original English version; we have performed Principal Components Analysis (PCA) (a dimension reduction technique). For current study Kaiser-Meyer-Olkin (KMO) test yielded an index of 0.764, suggesting a support for factor analysis. Bartlett’s test of sphericity is 985.518 and is highly significant (*P* = 000) (indicating a high probability of significant relationships between the variables).

### Principal Component Analysis (PCA)

Factor analysis is the most powerful statistical procedure for scrutinizing relations between observed and latent variables^10^. Three factors F1, F2, F3 viz “Perspective Taking”, “Compassionate Care” and “Standing in Patient’s Shoes” respectively emerged as shown in table no 6.

**Table 1:** Year of study and Gender distribution

| Year of Study (MBBS) | Male | Female | Total |
| --- | --- | --- | --- |
| 1<sup>st</sup> Year | 52 | 49 | 101 |
| 2<sup>nd</sup> Year | 60 | 42 | 102 |
| 3<sup>rd</sup> Year | 48 | 47 | 95 |
| 4<sup>th</sup> Year | 70 | 50 | 120 |
| Final Year | 66 | 37 | 103 |
| Total | 296 | 225 | 521 |

**Table 2:** Response Rate with respect to Gender and Year of Study

| Year of Study (MBBS) | Male | Female |
| --- | --- | --- |
| 1<sup>st</sup> Year | 48 | 45 |
| 2<sup>nd</sup> Year | 38 | 38 |
| 3<sup>rd</sup> Year | 39 | 38 |
| 4<sup>th</sup> Year | 32 | 43 |
| Final Year | 48 | 36 |
| Total | 205 | 200 |

**Table 3:** Mean Empathy Score with SD

|  | **<u>Item</u>** | **<u>Mean</u>**<br>**<u>(±SD)</u>** |
| --- | --- | --- |
| 1 | Physician's understanding of their patients' feelings and the feelings of their patients' families do not influence medical or surgical treatment. | **5.3 (2.05)** |
| 2 | Patients feel better when their physicians understand their feelings. | **6.6 (0.86)** |
| 3 | It is difficult for a physician to view things from a patient's perspective. | **3.9 (1.9)** |
| 4 | Understanding body language is as important as verbal communication in physician-patient relationship. | **6.1 (1.36)** |
| 5 | A physician's sense of humour results in a better clinical outcome. | **6.0 (1.37)** |
| 6 | Because patients are different, it is difficult to see things from patients' perspective. | **3.3 (1.77)** |
| 7 | Attention to patients' emotion is not important in history taking. | **5.3 (2.08)** |
| 8 | Attentiveness to patients' personal experience does not influence clinical outcome. | **5.2 (1.88)** |
| 9 | Physicians should try to stand in their patients' shoes when providing care to them. | **5.7 (1.77)** |
| 10 | Patients value a Physician's understanding of their feelings, which is therapeutic in its own right. | **6.2 (1.32)** |
| 11 | Patients' illness can only be cured by medical or surgical treatment; therefore, Physicians' emotional ties with their patients do not have a significant influence on medical or surgical treatment. | **5.2 (1.91)** |
| 12 | Asking patients what is happening in their personal lives is not helpful in understanding their physical complaints. | **5.5 (1.83)** |
| 13 | Physicians should try to understand what is going on in their patients' minds by paying attention to their non-verbal cues and their body language. | **6.0 (1.33)** |
| 14 | I believe that emotions have no place in the treatment of medical illness. | **5.3 (1.91)** |
| 15 | Empathy is a therapeutic skill without which the Physician's success is limited. | **5.9 (1.38)** |
| 16 | Physicians' understanding of the physical status of their patients, as well as that of their families is one important component of the Physician-patient relationship. | **5.8 (1.55)** |
| 17 | Physicians should try to think their patients in order to give better care. | **6.2 (1.17)** |
| 18 | Physicians should not allow themselves to be influenced by strong personal bonds between their patients and their family members. | **2.7 (1.89)** |
| 18 | I do not enjoy reading non-medical literature or the art. | **5.1 (2.06)** |
| 20 | I believe that empathy is an important therapeutic factor in medical treatment. | **6.2 (1.17)** |

**Table 4:** 3 Factor Mean Score with SD

|  |  |  |
| --- | --- | --- |
| Overall Empathy |  | 107.73 ±12.576 |
| Factor 1 | Patient's Perspective | 6.09 ±1.33 |
| Factor 2 | Compassionate Care | 4.97 ±1.96 |
| Factor 3 | Standing in patient's shoes | 3.63 ±1.85 |

**Table 5:** Cronbach’s Alpha if item deleted

|  |  |
| --- | --- |
| ItemNumber1 | .665 |
| ItemNumber2 | .673 |
| ItemNumber3 | .693 |
| ItemNumber4 | .684 |
| ItemNumber5 | .673 |
| ItemNumber6 | .688 |
| ItemNumber7 | .666 |
| ItemNumber8 | .664 |
| ItemNumber9 | .666 |
| ItemNumber10 | .669 |
| ItemNumber11 | .658 |
| ItemNumber12 | .670 |
| ItemNumber3 | .676 |
| ItemNumber14 | .650 |
| ItemNumber15 | .671 |
| ItemNumber16 | .657 |
| ItemNumber17 | .670 |
| ItemNumber18 | .697 |
| ItemNumber19 | .683 |
| ItemNumber20 | .667 |

**Table 6:**
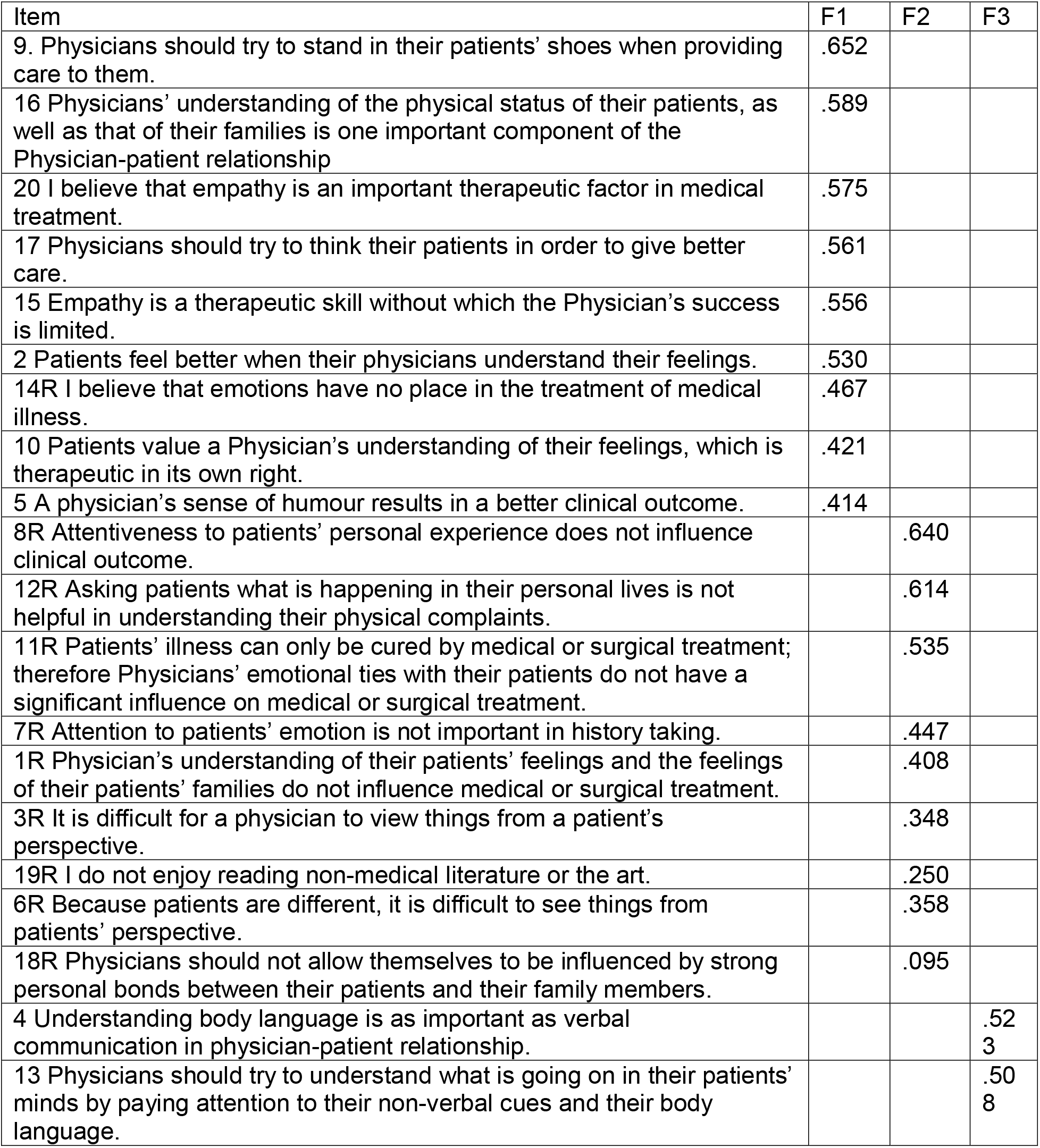
Factor analysis for Perspective Taking”, “Compassionate Care” and “Standing in Patient’s Shoes”

**Table 7.** Comparing the Mean scores (and standard deviation) of the three factors in the current and Italian Studies.

|  | <u>Current</u> | <u>Italian</u> |
| --- | --- | --- |
| <u>Overall empathy</u> | 107.73<br>+/-<br>12.576 | 108.71 ±<br>10.60 |
| <u>Factor 1</u><br>Patient's<br>Perspective | 6.09 +/-<br>1.33 | 5.56 ± 0.66 |
| <u>Factor 2</u><br>Compassionate<br>Care | 4.97 +/-<br>1.96 | 5.56 ± 0.66 |
| <u>Factor 3</u><br>Standing in<br>patient's shoes | 3.63 +/-<br>1.85 | 3.92 ± 1.24 |

## Discussion

The results of present study confirm the content, validity, reliability of Urdu version of JSE-S. Overall response rate for questionnaire was 77.88% which is comparatively less when compared to the study of Asma Mostafa et al who reported it to be 81.69%. Gender wise response rate was higher for female students (88.9%) as compared to male (69.26%). This is not surprising as literature shows that female exhibit higher response^12^, and are more compliant^13^; female scientists are known to be more collaborative than their male counterparts ^14,15^

The overall Cronbach’s alpha for present study was 0.684. Although a Cronbach alpha above 0.6 had been termed “acceptable” by some authors^16^, others^17^ prefer a figure exceeding 0.7. The deletion of only item 18 would raise Cronbach’s alpha to 0.7. This shows the reliability of the Urdu version of JSE-S.

The principal component analysis shows a three-factor solution that is somewhat similar to the pattern in other studies ^11,18,19^ This also provides support for the construct validity of the Urdu version of JSE-S.

It is not clear why our students have scored higher than the Italian students^19^ for Perspective taking and lower for Compassionate Care” and Standing in Patint’s shoes (Table 7). This is the first time that any study has shown such differences in three factors between two nations. Perspective taking reflects cognitive skills including information processing, reasoning, appraisal, and communicating empathy, as well as greater altruistic motivation^20^. Imagining the self and imagining the others in pain activate similar neural mechanisms; therefore enable one’s ability to perceive other’s pain in identical manner^21^. We found, higher ability among our students, to perceive others’ pain. This may be explained by the fact that Pakistan is badly affected, for more than 2 decade, by terrorism. As conditions of terrorism is improving since last 4-5 years it will be interesting to note if this trend continues in subsequent years.

Scoring lesser than their Italian compatriot in “Compassionate Care” and “Standing in Patient’s shoes” reflected lesser ability to address the patient’s psychosocial problems and understanding the patient’s experience, feelings and clues^19,20^. Currently study showed that Pakistani students find it difficult to address client’s psychosocial problems when compared to published literature ^22^. The reason for these differences is not clear but may be due to religious, cultural and economical issues. Islam discourages communication between men and women. In Pakistani culture too, such interaction is discouraged. Joshua Horden^23^ has mentioned that some of the religious and cultural taboos adversely affect communication with the patients. It is also known that the patients from higher social classes communicate more actively and show more affective expressiveness, eliciting more information from their doctor. Patients from lower social classes are often disadvantaged because of the doctor’s misperception of their desire and need for information and their ability to take part in the care process^24^. Hence the religion, culture and economical factors may have affected the empathy of our patients.

Finally, the item 18R “Physicians should not allow themselves to be influenced by strong personal bonds between their patients and their family members” has raised some concern. This is the only item where our students have scored very low (2.67 out of 7). This item has scored lowest in many other studies too^11,19^. Besides the fact that it is the only item whose deletion would raise Cronbach’s alpha to 0.7, lowering the overall internal consistency of the entire scale. It is also the only item that has failed to load significantly. The problem may lie in the phrasing/wording of the item. It perhaps suggests that it is asking about a non-professional, intimate relationship. The fact that it has been negatively worded may have added to the complexity of the item. I suggest that this item should be removed from JSE-S.

## Limitation of the study

This study has some limitations. Medical students who have participated in this survey belonged to Muhammad medical college only. Hence our sample is not representative of all Pakistani medical students. A replication of the study with a larger and more representative sample of Pakistani medical students will strengthen the confidence in the external validity and reliability of the findings. 406 out of 521 students have returned the form. It is possible that those who have taken interest may have more empathy than those 115 who have failed to return the form. The differences in groups may be attributed to cohort effects. Therefore, a study design using longitudinal annual assessment of the cohorts is warranted.

## Data Availability

Data associated with a paper is available at Muhammad Medical College, Mirpurkhas, and can be accessed by making an official request.

## Acknowledgments, Competing Interests

There is no competing interest or funding.

## References

1. Alcorta-GA, Gonzalez-GJ, Tavitas-HS, Rodrigues LF, Hojat M. Validity of the Jefferson Scale of Physician Empathy among Mexican medical students. Salud Ment. 2005. 28: 57-63.

2. Mohammadreza Hojat, Salvatore Mangione, Thomas J. Nasca, Mitchell J. M. Cohen et al. Measuring tool: Development and Preliminary Psychometric Data. Educational and Psychological Measurement; 2001. 61(2).

3. M. Hojat, J. S. Gonnella, S. Mangione, T. J. Nasca, M. Magee, “Physician empathy in medical education and practice: experience with the Jefferson scale of physician empathy,” Seminars in Integrative Medicine, vol. 1, no. 1, pp. 25-41, 2003.

4. Ward J, Schaal M, Sullivan J, Bowen ME, Erdmann JB, Hojat M. Reliability and validity of the Jefferson Scale of Empathy in undergraduate nursing students. J Nurs Meas. 2009;17(1):73-88.

5. Mohammadreza Hojat, Jennifer DeSantis, Stephen C. Shannon, Luke H, Mortensen et al. The Jeferson Scale of Empathy: a nationwide study of measurement properties, underlying components, latent variable structure, and national norms in medical students. Advances in Health Sciences Education (2018) 23:899-920 https://doi.org/10.1007/s10459-018-9839-9 1 3

6. https://www.jefferson.edu/university/skmc/research/research-medical-education/jefferson-scale-of-empathy.html accessed on 201141732011417320114173 at 201141732011417320114173

7. Hojat, M, LaNoue, M. Exploration and confirmation of the latent variable structure of the Jefferson scale of empathy. International Journal of Medical Education, 2014; 5:73-81. DOI: 10.5116/ijme.533f.0c41.

8. Kaiser, Henry F. “The Application of Electronic Computers to Factor Analysis.” Educational and Psychological Measurement, 1960: vol. 20, no. 1, Apr. 1960, pp. 141-151, doi: 10.1177/001316446002000116.

9. Brown, J. D. (1996). Testing in language programs Chapter 4. Upper Saddle River, NJ: Prentice Hall Regents, pp. 231-249.

10. Mohsen Tavakol, Reg Dennick. Making sense of Cronbach’s alpha. Int J Med Educ. 2011; 2: 53-55. doi: 10.5116/ijme.4dfb.8dfd.

11. Asma Mostafa, Rozina Hoque, Mohammad Mostafa, Md. Mashud Rana, Faisal Mostafa. Empathy in Undergraduate Medical Students of Bangladesh: Psychometric Analysis and Differences by Gender, Academic Year, and Specialty Preferences. ISRN Psychiatry Volume 2014, Article ID 375439, 7 pages http://dx.doi.org/10.1155/2014/375439.

12. Barbara Kastlungera, Stefan G. Dresslera, Erich Kirchlera, Luigi Mittoneb, Martin Voracek. Sex differences in tax compliance: Differentiating between demographic sex, gender-role orientation, and prenatal masculinization (2D:4D). Journal of Economic Psychology. 2010. 31(4), Pages 542-552.

13. Dimitrios G. Lyrakos The Impact of Stress, Social Support, Self-Efficacy and Coping on University Students, a Multicultural European Study. Psychology 2012. 3 (2), 143-149. http://dx.doi.org/10.4236/psych.2012.32022.

14. Hunter L, Leahey EE. Collaborative research in sociology: Trends and contributing factors. American Sociologist. 2008 Dec;39(4):290-306. https://doi.org/10.1007/s12108-008-9042-1.

15. B Ozel, H Kretschmer, T Kretschmer. Co-authorship pair distribution patterns by gender. Scientometrics 2014. 98 (1), 703-723.

16. Gong Feng-mei, MA Shi-hua, Tan Yong. Empirical Study on the Influence between Logistics Information Capabilities and Supply Chain Performance. Industrial Engineering and Management. 2007-02.

17. Nunnally, J. C., & Bernstein, I. H. (1994). Psychometric theory (3rd ed.). New York: McGraw-Hill. Google Scholar.

18. Franck Zenasni, Emilie Boujut, Aude Woerner and Serge Sultan. Burnout and empathy in primary care: three hypotheses. British Journal of General Practice 2012; 62 (600): 346-347. DOI: https://doi.org/10.3399/bjgp12X652193.

19. Leombruni, P, Di Lillo, M., Miniotti, M. Angelo Picardi, Guido Alessandri et al. Measurement properties and confirmatory factor analysis of the Jefferson Scale of Empathy in Italian medical students. Perspect Med Educ; 2014. 3, 419-430. https://doi.org/10.1007/s40037-014-0137-9.

20. Hojat M: Empathy in patient care: Antecedents, development, measurement, and outcomes. Page No 80. New York, Springer, 2007.

21. Claus Lamm, C. Daniel Batson and Jean Decety. The Neural Substrate of Human Empathy: Effects of Perspective-taking and Cognitive Appraisal. Journal of Cognitive Neuroscience. 2007: 19(1) 42-58.

22. Mohsen Tavakol and Reg Dennick. Making sense of Cronbach’s alpha. Int J Med Educ. 2011; 2: 53-55.

23. Joshua Hordern. Ethics and Communication Skills. Medicine. 2016. 44 (10), P589-592, DOI: https://doi.org/10.10167j.mpmed.2016.07.011.

24. S. Willems, S. De Maesschalck, M. Deveugele, A. Derese J, De Maeseneer. Socio-economic status of the patient and doctor-patient communication: does it make a difference? Patient Education and Counseling. 2005. 56 (2). Pages 139-146.

